# Is anxiety a pathway to Alcohol Use Disorders? A phenome-wide association study of the *GABRA2* coding variant rs279858

**DOI:** 10.1101/2022.11.21.22282301

**Authors:** Alison K. Merikangas, Rachel L. Kember, Martin H. Plawecki, Chella Kamarajan, Grace Chan, Lance Bauer, Jacquelyn L. Meyers, John I. Nurnberger, John Kramer, Bernice Porjesz, Howard J. Edenberg, Laura Almasy

## Abstract

Alcohol use disorders (AUDs) and related electrophysiological endophenotypes have been associated with the *GABRA2* gene. However, the causal variants in *GABRA2* and their mechanisms of influence on AUD and its correlates have not been established. Here we investigate the phenotypic spectrum of a *GABRA2* coding variant (rs279858) through a phenome-wide association study (PheWAS) in two open-source datasets.

We applied the PheWAS approach to identify a broad range of phenotypes associated with rs279858 in the MRC IEU OpenGWAS PheWAS and the Open Targets Genetics Portal. These databases extend the array of phenotypes beyond those available in electronic health records (EHR) to include numerous non-medical phenotypes and traits. We then followed up the results from those exploratory associations by examining the genetic correlations between our “top hits” and alcohol- and smoking-related phenotypes.

In both data sources, rs279858 (C effect allele) was associated with anxiety-related phenotypes, including reduced risk-taking behavior and an increase in nervous feelings, as well as reduced number of lifetime sexual partners. Follow-up analyses revealed that these phenotypes were genetically correlated with each other and with alcohol- and smoking-related phenotypes.

This work illustrates the utility of the PheWAS approach, particularly for phenotypes that extend beyond those that are typically captured in EHR data. In fact, the associations described here are all behavioral rather than clinical phenotypes. We postulate that these traits may be related to anxiety or behavioral inhibition that has been identified as a risk factor for AUD, and may represent pathophysiological intermediaries between *GABRA2* and AUD.

## Introduction

Alcohol Use Disorder (AUD) is a complex condition defined by a pattern of unhealthy, problematic use of alcohol with significant clinical impairment. There are both genetic and environmental contributions to its development. AUD has considerable public health, social, and economic costs with an estimated 5.9% of people meeting past-year criteria [1] and 13.8% in the United States meeting criteria for lifetime AUD [2]. Despite a reported heritability of nearly 50%, and substantial effort to establish the underlying genetic etiology, few genes aside from the alcohol metabolizing genes *ADH1B* and *ALDH2* have been consistently associated with AUD in population-based genome-wide association studies (GWAS)[3]. However, a recent large meta-analysis including 435,563 individuals of European descent has now identified 29 associated loci for ‘problematic alcohol use’ (PAU)[4].

Both linkage and association studies of samples enriched for AUD have implicated the gene encoding gamma-aminobutyric acid type A receptor subunit alpha2 (*GABRA2*) in the etiology of alcohol use disorder [5-8]. The gamma-aminobutyric acid receptor subunit alpha-2 protein is a subunit of the GABA-A receptor. RNA-seq studies have demonstrated higher *GABRA2* expression in brain, kidney, and small intestine as comparted to other body tissues [9,10]. *GABRA2* has also been implicated in EEG changes in those with AUD, especially the beta rhythm of the resting EEG [5,11,12]. *GABRA2* may act via neuromodulation since it has been associated with both AUD and EEG phenotypes [5].

In addition to AUD, associations have been reported between *GABRA2* and several substance misuse phenotypes, including vulnerability to both nicotine dependence (ND) and cannabis dependence (CD)[13] as well as subjective response to alcohol and other alcohol-related reward processes [14,15]. *GABRA2* has also been implicated in the disrupted structural connectome of reward circuits in heroin abusers [16], and in other neuropsychiatric-related phenotypes, including epilepsy [17], age at onset of Alzheimer’s disease [18], and risk-taking behavior [19], as well as antipsychotic-associated weight gain [20].

Though multiple single nucleotide polymorphisms (SNPs) within *GABRA2* have been associated with both alcohol use and electrophysiological phenotypes, multiple studies have shown the strongest association signal for rs279858 and SNPs that are in linkage disequilibrium (LD) with this lead variant [5,21]. The rs279858 SNP in *GABRA2* is one of the few loci from the candidate gene analysis era that was associated with alcohol dependence in the Study of Alcohol Addiction: Genetics and Addiction (SAGE)[8]. Specifically, G/G carriers appear to be more prone to AUD than A/A or A/G carriers [22,23]. However, in some publications, genotypes were called from the complementary DNA strand with alleles C and T, in which case the C allele is the one associated with higher risk. A functional role for this synonymous change in rs279858 has not been established. Researchers have postulated that its actions involve the modulation of both cognition and reward systems [16]. However, this SNP has not been associated with AUD in the largest case-control GWAS of AUD [4] and PAU [24] to date, but it has been implicated in phenotypic manifestations of alcohol use in some race/ethnicity subgroups [25] and in sex-specific analyses [26].

In parallel to the GWAS approach that attempts to identify markers that may influence the risk of AUD across the genome, phenome-wide association studies (PheWAS) are employed to identify a range of phenotypes that may be associated with a particular genetic variant. PheWAS test the association between a genetic variant (e.g., a SNP, gene, or polygenic score), and an array of phenotypes that range from International Classification of Diseases (ICD) codes in electronic health records (EHR)[27,28] to data from epidemiologic studies and biobanks that have been supplemented with questionnaires that assess lifestyle and behavioral traits [29]. PheWAS can identify associations of risk loci with novel traits that have not been previously linked to AUD. This data-driven approach can highlight numerous additional traits or conditions that may be associated with a genetic variant that might not have been investigated in analyses driven by known relationships among phenotypes. The PheWAS approach, e.g., PheWAS of polygenic scores [30,31] and other AUD-associated variants [32] has also been employed to elucidate comorbid conditions that may also contribute to the developmental trajectory of AUD.

Here we apply the PheWAS approach to several open-source tools in order to examine the phenotypic spectrum of an AUD-associated *GABRA2* coding variant (rs279858) that may provide indices of potential causal systems and pathways. We employ three approaches to address the exploratory nature of this work including: a) correcting for multiple comparisons; b) conducting a “pseudo” replication by using two curated databases containing multiple large studies; and c) inspecting the genetic correlations within the identified phenotypes and between these phenotypes and alcohol-related phenotypes.

## Materials and Methods

### PheWAS data sources

The PheWAS approach [27] was used to investigate the range of phenotypes associated with rs279858 using two online tools; the Medical Research Council (MRC) Integrative Epidemiology Unit (IEU) at the University of Bristol OpenGWAS PheWAS tool [33] (https://gwas.mrcieu.ac.uk/phewas/), and the Open Targets Genetics Portal [34] (https://genetics.opentargets.org/). Supplementary Table 3 lists the included data sources for both tools.

The MRC IEU OpenGWAS PheWAS queries a manually curated collection of eighteen batches of GWAS summary datasets encompassing over 14,000 individual GWAS that includes the UK Biobank (UKBB) and other national databases such as the Japanese Biobank [35] (https://biobankjp.org/en/), and FinnGen (https://www.finngen.fi/en), as well as the National Human Genome Research Institute (NHGRI)-European Bioinformatics Institute (EBI) GWAS Catalog summary statistics database. We searched rs279895 using their tool (https://gwas.mrcieu.ac.uk/phewas/), which only reports results filtered to p≤0.001, and then used the *ieugwasr* R package (R version 4.0.4) to connect to the application programming interface (API) and downloaded the complete results file (version 3.8.5).

The Open Targets Genetics Portal includes data from FinnGen (https://www.finngen.fi/en), the National Human Genome Research Institute (NHGRI)-European Bioinformatics Institute (EBI) GWAS Catalog summary statistics database [36] (GWAS Catalog, https://www.ebi.ac.uk/gwas/), and UKBB (http://www.nealelab.is/uk-biobank). We searched rs279858 on the Open Targets Genetics site (https://genetics.opentargets.org/, version 22.02) which maps this SNP to its location GRCh38: 4:46,312,576 (GRCh37: 4:46,314,593, https://genetics.opentargets.org/variant/4_46312576_T_C). Only traits associated at p<0.005 are returned and used for analysis.

### PheWAS statistical significance

Some traits that have been examined in independent GWAS analyses of the biobanks have also been deposited in the GWAS Catalog. Therefore, traits may appear in both the OpenGWAS and Open Targets results, and on occasion multiple times in each. For such situations, we opted to retain the results from the instance with the smallest p-value. We adjusted the p-value to correct for the 16482 traits that appeared in the downloadable OpenGWAS dataset, which resulted in a Bonferroni-corrected significance threshold for this study of p<4.00×10^−6^. The Open Targets tool provides a Bonferroni-corrected phenome-wide significance threshold of p<9.77×10^−5^. The overlap in contributing data sources in shown in Supplementary Table 3.

### SNP-based heritability

In order to determine the likelihood of detecting genetic signals for alcohol-related phenotypes in the UKBB, we examined the SNP-based heritability (*h*^2^) in the UKBB generated by the Neale Lab [37]. They provide an automated calculation of SNP-heritability via partitioned linkage disequilibrium score regression (LDSC)[38], with *h*^2^ values using a liability scale for binary phenotypes. We used this information to gauge the overall genetic signal for UKBB alcohol-related phenotypes queried in our PheWAS analysis, by examining the heritability and statistical significance of component items of the Alcohol Use Disorders Identification Test (AUDIT)[39] present in the UKBB.

### Genetic correlation

We used results generated by the Neale Lab on the genetic correlation (*r*_g_) between phenotypes in the UKBB via LDSC (https://ukbb-rg.hail.is/) to explore unexpected results from the PheWAS. Statistically significant phenotypes from the PheWAS analyses were compared to one another, and to the available alcohol, tobacco, and cannabis use phenotypes. This information can be used to evaluate whether the identified phenotype may be indexing another trait that does not demonstrate statistical significance due to sample size. A Bonferroni corrected p-value of 2.92×10^−4^ was employed to correct for the number of correlations that we examined across the selected traits (171 pairs).

## Results

The PheWAS results at p<0.005 from both the MRC IEU OpenGWAS and the Open Targets PheWAS are shown in Supplementary Table 1 and 2, respectively. Supplementary Table 3 lists the data sources and highlights their overlapping and independent contributions to the results presented here.

### MRC IEU OpenGWAS

The strongest associations for rs279858 (C effect allele) were with reduced risk-taking behavior, an increase in nervous feelings, reduced number of lifetime sexual partners, and reduced length of mobile phone use. Table 1 shows the top ten associations of rs279858 in the IEU analysis. For traits that appeared more than once, the instance with the smallest p-value was retained. Figure 1a and Supplementary Table 1 show the complete results at p<0.005.

**Table 1:** Top ten associations of rs279858 in the OpenGWAS Analysis.

| Trait | Data Source | Total N | Effect Size | p-value |
| --- | --- | --- | --- | --- |
| **Risk taking** | IEU UKBB | 446,279 | **-0.0052** | **2.10×10<sup>-08</sup>** |
| **Nervous feelings** | IEU UKBB | 450,700 | **0.0045** | **4.60×10<sup>-07</sup>** |
| **Lifetime number of sexual partners** | IEU UKBB | 378,882 | **-0.0090** | **1.10×10<sup>-06</sup>** |
| **Length of mobile phone use** | IEU UKBB | 456,972 | **-0.0134** | **2.80×10<sup>-06</sup>** |
| Natural Killer T Absolute Count | EBI Database | 3,653 | 0.1123 | 1.02×10<sup>-05</sup> |
| HLA DR on HLA DR+ T cell | EBI Database | 3,060 | -0.1071 | 4.10×10<sup>-05</sup> |
| Natural Killer T %T cell | EBI Database | 3,669 | 0.1046 | 4.34×10<sup>-05</sup> |
| Natural Killer T %lymphocyte | EBI Database | 3,669 | 0.1020 | 7.47×10<sup>-05</sup> |
| Milk type used: Never/rarely have milk | Neale UKBB | 360,806 | -0.0017 | 8.08×10<sup>-05</sup> |
| MAP kinase-activated protein kinase 2 | Sun et al. 2018 [40] | 3,301 | 0.0941 | 1.35×10<sup>-04</sup> |

Statistically significant findings are highlighted in bold font. IEU=Integrative Epidemiology Unit; EBI=European Bioinformatics Institute; Sun et al. 2018 [40] is an independent study included in the OpenGWAS database

**Figure 1:**
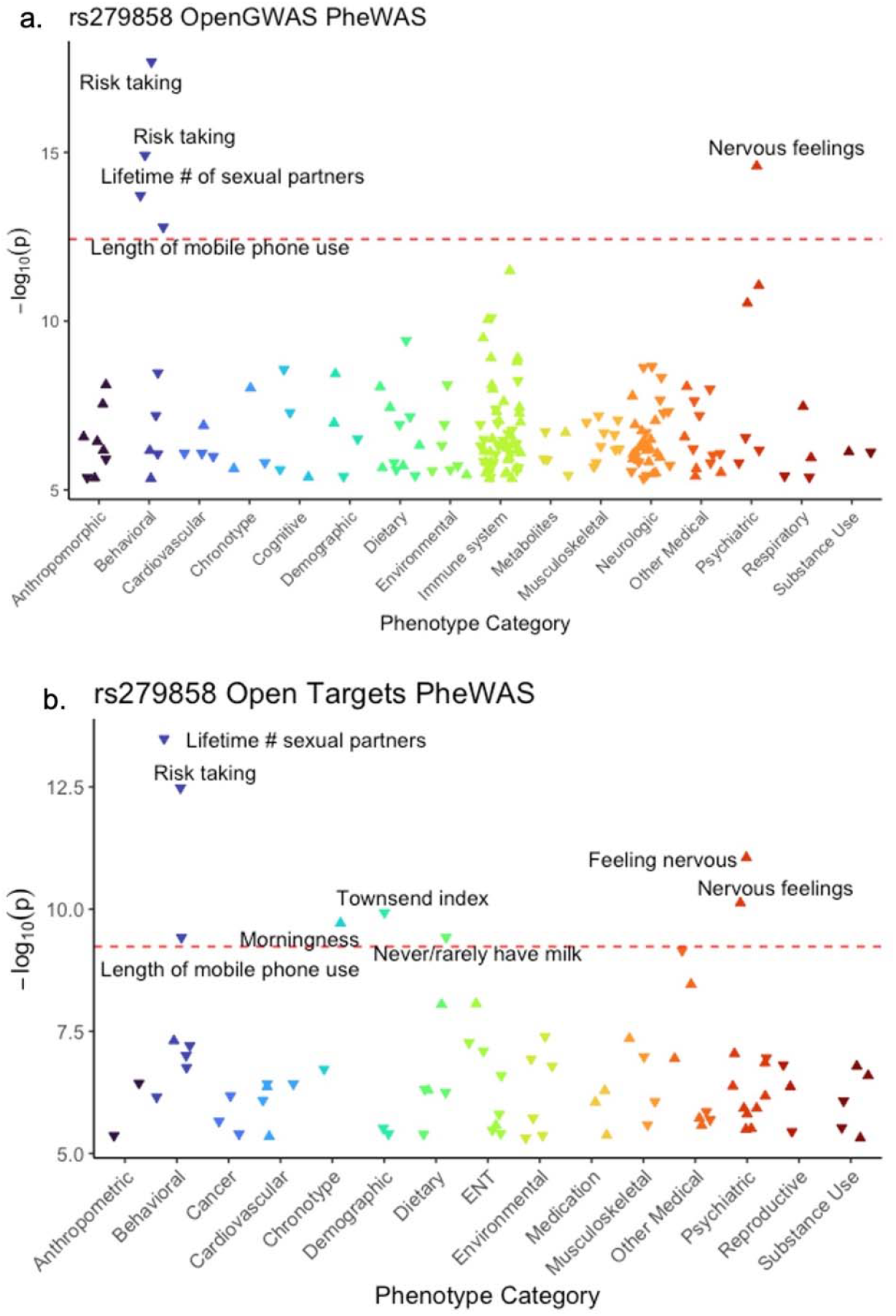
OpenGWAS and Open Targets rs279858 PheWAS plots. Phenome-wide association study results for rs279858 from the OpenGWAS PheWAS tool (a) and the Open Targets PheWAS tool (b). Traits with a p-value p<0.005 are shown, with the down arrow representing a negative beta, and the up arrow representing a positive beta (C effect allele). In (a) the dashed red line represents a Bonferroni-corrected phenome-wide significance threshold of p<4.00×10^−6^. In (b) the dashed red line represents a Bonferroni-corrected phenome-wide significance threshold of p<9.77×10^−5^

### Open Targets Genetics

In the Open Targets analysis, rs279858 (C effect allele) was associated with a reduced level of self-reported number of lifetime sexual partners, reduced risk-taking behavior, an increase in nervous feelings, a lower Townsend deprivation index at recruitment (a measure of socioeconomic status within a population encompassing employment, car and home ownership, and household overcrowding by post code)[41], getting up earlier in the morning, never/rarely having milk, and a reduced length of mobile phone use. Table 2 shows the top ten associations for rs279858 in the Open Targets analysis. Figure 1b and Supplementary Table 2 show the complete results at p<0.005. No alcohol, tobacco, or other substance use phenotypes were among the top rs279858 associations in either PheWAS.

**Table 2:** Top ten associations of rs279858 in the Open Targets Analysis.

| Trait | Data Source | Total N | Effect Size | p-value |
| --- | --- | --- | --- | --- |
| **Lifetime number of sexual partners** | **Neale UKBB** | **296609** | **-0.0098** | **$1.39 \times 10^{-06}$** |
| **Risk taking** | **Neale UKBB** | **348549** | **-0.0250** | **$3.80 \times 10^{-06}$** |
| **Nervous feelings** | **Nagel et al. 2018 [42]** | **373121** | **0.0101** | **$1.58 \times 10^{-05}$** |
| **Townsend deprivation index at recruitment** | **Neale UKBB** | **360763** | **-0.0283** | **$4.87 \times 10^{-05}$** |
| **Getting up in morning (Morningness)** | **Neale UKBB** | **360231** | **0.0072** | **$6.04 \times 10^{-05}$** |
| **Never/rarely have milk \| milk type used** | **Neale UKBB** | **360806** | **-0.0531** | **$8.08 \times 10^{-05}$** |
| **Length of mobile phone use** | **Neale UKBB** | **356618** | **-0.0125** | **$8.11 \times 10^{-05}$** |
| Renal/kidney failure \| non-cancer, self-reported | Neale UKBB | 361141 | 0.4043 | $2.12 \times 10^{-04}$ |
| Both eyes \| which eye(s) affected by astigmatism | Neale UKBB | 9749 | 0.1130 | $3.15 \times 10^{-04}$ |
| Oat cereal \| cereal type | Neale UKBB | 299898 | 0.0214 | $3.19 \times 10^{-04}$ |

Statistically significant findings are highlighted in bold font. Nagel et al. 2018 [42] is an independent study included in the Open Targets database

### SNP-based heritability of AUDIT phenotypes in UKBB

Table 3 shows the AUDIT items in the UKBB, their SNP-based heritability, associated p-value, and the sample size for each item. The final column shows the p-value that was generated for that item in our MRC IEU OpenGWAS PheWAS analysis. The AUDIT items that showed statistically significant SNP-based heritability included: “Frequency of drinking alcohol”; “Amount of alcohol drunk on a typical drinking day”; and “Frequency of consuming six or more units of alcohol”. Notably, not all of the AUDIT items were included in the PheWAS, and the ones that were included were not associated with rs279858.

**Table 3:**
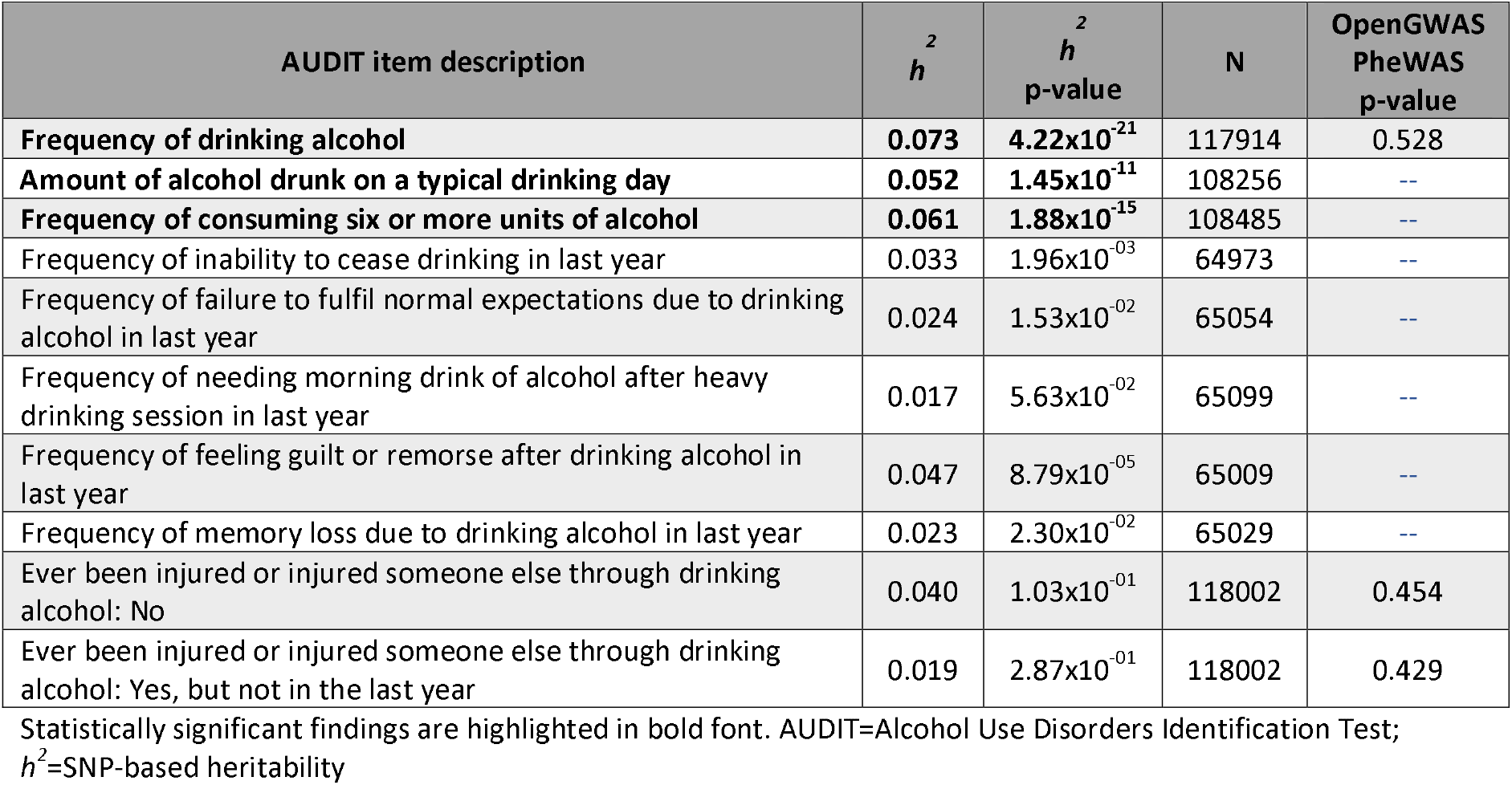
Heritability of alcohol phenotypes in the UKBB from the MRC IEW OpenGWAS PheWAS

### Genetic correlation

The genetic correlations between the statistically significant UKBB phenotypes from the PheWAS, and UKBB alcohol, tobacco, and substance use phenotypes are shown in Figure 2 (also see the full results in Supplementary Table 4). After a conservative Bonferroni correction (p<2.92×10^−4^), statistically significant genetic correlations were observed among the majority of the PheWAS phenotypes with the exception of the dietary phenotypes [i.e., “Milk type used: Never/rarely have milk” and “Cereal type: Oat cereal (e.g., Ready Brek, porridge)]”, which were nominally correlated with the other PheWAS traits (p<0.05) but did not survive correction for multiple testing. The top hits from the PheWAS were also genetically correlated with most of the smoking and alcohol phenotypes at a standard p<0.05. However, “Getting up in morning”, “Nervous feelings”, and the dietary phenotypes were not genetically correlated with smoking after employing correction for multiple comparisons. Similarly, fewer of the top hits were genetically correlated with alcohol phenotypes after control for multiple comparisons. Notably, “nervous feelings” was not genetically correlated with alcohol use. In addition, risk-taking and lifetime number of sexual partners were genetically correlated with being a previous drinker.

**Figure 2:**
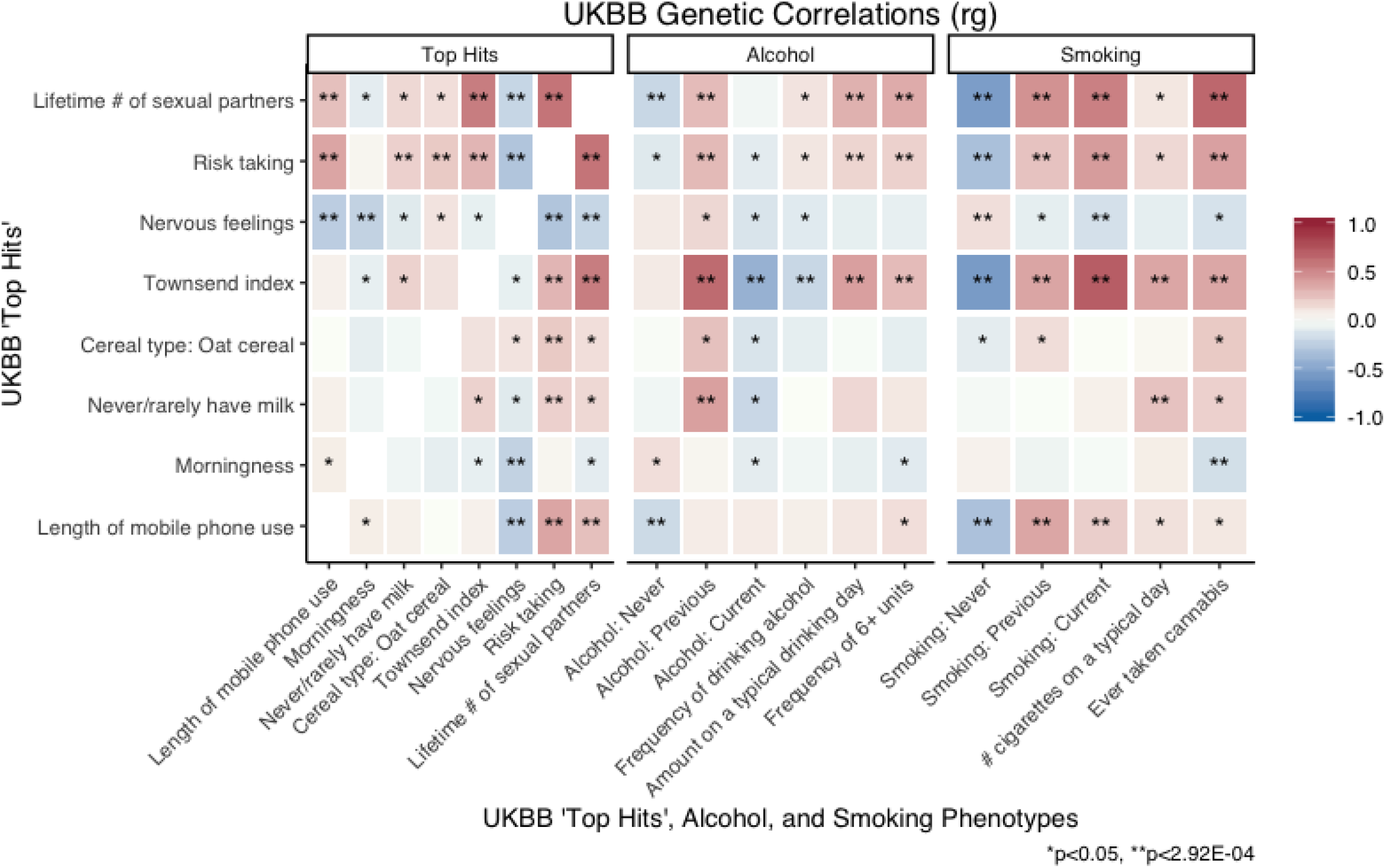
Genetic correlations among top UKBB rs279858 PheWAS hits, alcohol, and smoking phenotypes. The genetic correlations between the top rs279858-associated phenotypes, alcohol, and smoking phenotypes in the UKBB are shown. A single asterisk represents a nominally significant p-value p<0.05, and a double asterisk represents a p-value that meets the Bonferroni corrected threshold of p-value p<2.92×10^−4^. The darker blue represents a negative correlation, while the darker red represents a positive correlation.

## Discussion

In both open-source PheWAS analyses, the C allele of rs279858, the top AUD SNP within a *GABRA2* LD block, was associated with an increase in nervous feelings, a reduced level of self-reported risk-taking behavior, reduced number of lifetime sexual partners, and shorter duration of mobile phone use. We postulate that these traits may be related to reduced novelty seeking/behavioral inhibition or anxiety that has been identified as a risk factor for alcohol misuse [43,44]. These phenotypes have been shown to underlie anxiety-related symptoms and disorders that have been involved in pathways to AUD [45]. Our analyses of the genetic correlations both within these the anxiety/inhibition phenotypes as well with a large number of alcohol related behaviors suggests that the phenotypic links may share overall genetic architecture with AUD in addition to their common association with rs279858.

This work builds upon the substantial body of research that has attempted to identify genetic and environmental risk factors that may inform the multifactorial architecture of AUD. The broad range of phenotypes examined in these PheWAS tools, and particularly in the UKBB, extend far beyond those that are typically captured in EHR data. It is also notable that the effect sizes for the behavioral traits are quite small (∼0.005-0.05), which may explain why these associations are only seen in large samples.

The strongest associations that survived correction for multiple comparisons were for anxiety phenotypes and reduced novelty seeking/behavioral inhibition. The association between rs279858 C allele and the anxiety phenotypes in the PheWAS confirms the links between AUD and anxiety disorders, particularly social anxiety, reported in earlier family [46,47] and twin studies [48]. Furthermore, specific anxiety disorder subtypes have been shown to elevate the risk of subsequent onset of both alcohol use and dependence in community surveys worldwide [2,49]. The heterogeneity and complexity of this association has been described in several earlier reviews [50,51]. The convergence of our PheWAS findings with these genetic epidemiologic studies suggests that inquiry into potential mechanisms for the role of genetic factors indexed by this SNP may inform this potential pathway to AUD.

Our finding of the associations between anxiety related phenotypes with the C allele of rs279858 is consistent with reports of an association between externalizing behavior, including substance use and greater risk taking, with the opposite T allele of rs279858 allele [52-56]. In contrast, direct tests of association show that the C allele is associated with AUD [5,21,52]. Analyses of an adolescent sample [52] revealed indirect associations of AUD with the T allele through externalizing factors and direct associations of problematic alcohol use with the C allele. This suggests that the role of *GABRA2* in AUD may be multi-faceted and developmentally expressed. In the large nationally representative sample of adults in the National Comorbidity Survey [57], anxiety and externalizing were each independently associated with higher risk of AUD, but there was a potential interactive effect between anxiety and externalizing disorders that was associated with reduced rates of AUD.

The lack of an association between rs279858 with alcohol traits assessed in the UKBB [(e.g., consumption measures (alcohol intake frequency, p=0.69) and self-reported alcohol use problems (ever addicted to alcohol, p=0.04)] was not surprising given the lack of previous associations between rs279858 and AUD phenotypes in general population-based samples that were not recruited for substance use disorder studies. However, these analyses do show strong genetic correlations between our top PheWAS hits and alcohol and smoking phenotypes in UKBB. A potential explanation may be that the association of alcohol phenotypes with *GABRA2* has been found in samples enriched for AUD but not in a volunteer sample of older adults such as the UKBB, which is predominantly female and among which there was a low frequency of AUD. Another consideration is that a smaller proportion of participants completed some of the alcohol use modules in the UKBB, meaning that these traits have smaller sample sizes than the ones identified in our PheWAS.

*GABRA2* has been associated with a number of other traits in GWAS, including epilepsy [17,58], post bronchodilator FEV1/FVC ratio [59], Alzheimer disease and age of onset [18], Mononucleosis [60], general cognitive ability [61], general risk tolerance (MTAG) [19], adventurousness [19], protein quantitative trait loci (liver)[62], and Type 1 diabetes (age at diagnosis)[63]. It is notable that the association findings for general cognitive ability, general risk tolerance, and adventurousness were found in UKBB data. These traits are composite phenotypes formed from the UKBB traits identified in our PheWAS that include nervous feelings and reduced risk-taking and lend credence to the theory that the impact of *GABRA2* on alcohol-related phenotypes is through anxiety or risk taking.

The effects of the *GABRA2* gene in basic animal model studies provide intriguing parallels to our findings. Numerous phenotypes that overlap our association findings have been found in *GABRA2* knockout mice [64] (http://www.informatics.jax.org/marker/phenotypes/MGI:95614). In particular, *GABRA2* has been shown to impact parallel behavioral phenotypes in mice, including increased anxiety-related response and an abnormal conditioned emotional response, as well as medical phenotypes in the immune and nervous systems (Supplementary Table 5). Moreover, *GABRA2* has been related to hypersensitivity to the acute effects of alcohol [65] and is a binding site for benzodiazepines known to decrease anxiety [66]. There is further indirect evidence that neuroactive steroids may be involved in some of the subjective effects of alcohol reflecting the role of this SNP on both immune function and alcohol and benzodiazepine effects and metabolism [25,67-69]. It would be interesting to examine the impact of this marker on the preliminary evidence for the efficacy of gabapentin, a gamma aminobutyric acid (GABA) analogue, in the treatment of AUD [70].

### Strengths & limitations

The main strength of this study is the application of the PheWAS approach to identify potential influences of the *GABRA2* gene that extend beyond the diagnostic code-based EHR. We include a wide range of other biological and social phenotypes that may reflect the multifactorial etiology of AUD.

This work has several limitations that should be considered in interpreting these findings. First, phenotypes can appear multiple times in the same database or in different databases. This leads to a lack of independence in the analyses, which creates a challenge for appropriately adjusting for multiple comparisons, meaning the Bonferroni method is extremely conservative in this case. Second, the wide range of phenotypes that extend beyond medically relevant conditions to include behavior, temperamental traits, and social variables may include variables that are neither empirical or conceptual correlates of AUD. This may make it difficult to interpret the results in the context of AUD, as exemplified by the associations with dietary phenotypes. Third, there is overlap in the two databases that were used for these analyses. Although the use of the UKBB data is a strength of this study, it is also a limitation because these findings may have overshadowed those of the smaller studies. Fourth, the applicability of this approach for understanding mechanisms of AUD relies on the relevance of the *GABRA2* gene, which although shown in both enriched samples and basic animal model studies, has not been identified in the large GWAS studies of AUD. Nevertheless, the similarity in the phenotypic findings from the PheWAS and the knockout mouse studies suggest that we may be honing in on relevant phenotypic manifestations of this gene.

## Conclusion

In conclusion, we have shown that rs279858 (C effect allele) is associated with anxiety phenotypes, which may represent pathophysiological intermediaries between *GABRA2* and AUD. If replicated, these findings would confirm the utility of identifying anxiety as a target of prevention and intervention of AUD. Future in depth investigation of the potential genetic and biologic mechanisms for the link between this genetic system and its correlates in the development of AUD is warranted.

## Data Availability

All data produced in the present study are available upon reasonable request to the authors.

## Acknowledgments

The support of Hannah L. Bocchinfuso, Resource Coordinator for the Almasy Lab, is invaluable. We are indebted to all of the researchers who deposit their data into central repositories, and those who developed the online tools used to complete this research.

The Collaborative Study on the Genetics of Alcoholism (COGA), Principal Investigators B. Porjesz, V. Hesselbrock, T. Foroud; Scientific Director, A. Agrawal; Translational Director, D. Dick, includes eleven different centers: University of Connecticut (V. Hesselbrock); Indiana University (H.J. Edenberg, T. Foroud, Y. Liu, M. Plawecki); University of Iowa Carver College of Medicine (S. Kuperman, J. Kramer); SUNY Downstate Health Sciences University (B. Porjesz, J. Meyers, C. Kamarajan, A. Pandey); Washington University in St. Louis (L. Bierut, J. Rice, K. Bucholz, A. Agrawal); University of California at San Diego (M. Schuckit); Rutgers University (J. Tischfield, R. Hart, J. Salvatore); The Children’s Hospital of Philadelphia, University of Pennsylvania (L. Almasy); Virginia Commonwealth University (D. Dick); Icahn School of Medicine at Mount Sinai (A. Goate, P. Slesinger); and Howard University (D. Scott). Other COGA collaborators include: L. Bauer (University of Connecticut); J. Nurnberger Jr., L. Wetherill, X., Xuei, D. Lai, S. O’Connor, (Indiana University); G. Chan (University of Iowa; University of Connecticut); D.B. Chorlian, J. Zhang, P. Barr, S. Kinreich, G. Pandey (SUNY Downstate); N. Mullins (Icahn School of Medicine at Mount Sinai); A. Anokhin, S. Hartz, E. Johnson, V. McCutcheon, S. Saccone (Washington University); J. Moore, Z. Pang, S. Kuo (Rutgers University); A. Merikangas (The Children’s Hospital of Philadelphia and University of Pennsylvania); F. Aliev (Virginia Commonwealth University); H. Chin and A. Parsian are the NIAAA Staff Collaborators. We continue to be inspired by our memories of Henri Begleiter and Theodore Reich, founding PI and Co-PI of COGA, and also owe a debt of gratitude to other past organizers of COGA, including Ting-Kai Li, P. Michael Conneally, Raymond Crowe, and Wendy Reich, for their critical contributions. This national collaborative study is supported by NIH Grant U10AA008401 from the National Institute on Alcohol Abuse and Alcoholism (NIAAA) and the National Institute on Drug Abuse (NIDA).

## Author Contributions

Conceptualized study: AKM, LA; Conducted analyses: AKM, RLK; Drafted manuscript: AKM, LA; Reviewed and edited manuscript: LA, LB, CG, HJE, CK, RLK, JK, AKM, JLM, JN, MHP, BP.

## Supplemental Material

Table S1: OpenGWAS rs279858 PheWAS results for p<0.005

Table S2: Open Targets rs279858 PheWAS results for p<0.005

Table S3: OpenGWAS and Open Targets PheWAS data sources

Table S4: Genetic correlations between the statistically significant UKBB phenotypes from the PheWAS, and UKBB alcohol, tobacco, and substance use phenotypes

Table S5: *GABRA2* Mammalian Phenotype Ontology Annotations (MGI:95614)

